# Diagnostic Value of Genetic Mutation Analysis and Mutation Profiling of cfDNA in Intraocular Fluid for Vitreoretinal Lymphoma

**DOI:** 10.1101/2021.10.18.21265102

**Authors:** Xiaoqing Chen, Yunwei Hu, Wenru Su, Shizhao Yang, Xiaoxiao Wang, Ping Zhang, Xiaoyu Hong, Chuqiao Liang, Zhuyun Qian, Ziqiang Li, Yong Tao, Huiqiang Huang, Dan Liang

## Abstract

**Objective:** Vitreoretinal lymphoma (VRL) is a rare but aggressive masquerade syndrome, with challenging diagnosis. Since the tumor-related genetic mutation analysis based on cell-free DNA (cfDNA) are underutilized in ocular oncology, we aimed to test the VRL diagnostic value of cfDNA genetic mutation analysis using intraocular fluid (IOF) samples and to identify its mutation profile.

**Subjects:** Seventeen VRL and 6 uveitis patients from Zhongshan Ophthalmic Centre were selected as training group, and 5 VRL and 5 uveitis patients from Beijing Chaoyang Hospital were selected as validation group.

**Methods:** The medical records and genetic mutation analysis using a panel containing 446 tumor-related genes of included patients were retrospectively reviewed. We analyzed the mutation profile, and identified the molecular subtypes and subdivisions of B-cell differentiation pathways of our VRL patients.

**Main outcomes:** The cfDNA genetic mutations detected in IOF.

**Results:** The VRL patients from the training group exhibited abundant cfDNA mutations in IOF (sensitivity 100%), while only 2 out of 6 uveitis patients were positive (specificity 67%). However, the number of cfDNA mutations observed in VRL patients was higher than that of uveitis patients. The mutation-positive patients from the validation group were diagnosed with VRL while the mutation-negative patients were diagnosed with uveitis (sensitivity and specificity 100%). VRL patients were characterized by the high mutation frequencies of *PIM1, MYD88, CD79B, ETV6*, and *IRF4*.

**Conclusions:** The genetic mutation analysis of IOF samples represents a feasible diagnosis method for VRL with 100% sensitivity; it could track genetic profiles; thus, revealing genetic heterogeneity of VRL.

**Statement of translational relevance:** The anterior chamber paracentesis and diagnostic vitrectomy have been widely used in ophthalmic clinics, as they are less invasive sampling techniques for liquid biopsies. Genetic mutation analysis of cell-free DNA (cfDNA) of intraocular fluid using a panel containing 446 targeted genes represents a feasible method with 100% sensitivity for vitreoretinal lymphoma (VRL) diagnosis. Furthermore, it can address cytological diagnostic issues including limited cellular yield, cell lysis associated with the fragile nature of lymphoma cells, and high risks associated with retinal tissue biopsies. Furthermore, it allows for genetic profile tracking, having the potential to reveal genetic heterogeneity and molecular characteristics of VRL in the future

## Introduction

Vitreoretinal lymphoma (VRL) is a rare intraocular malignant lymphoma affecting the vitreous and/or the retina that can occur as a primary lymphoma or as a secondary manifestation of a primary central nervous system (CNS) lymphoma^1^. The vast majority of cases are histopathologically classified as diffuse large B-cell lymphoma (DLBCL)^2^.

Clinically, VRL masquerades posterior- and pan-uveitis, as it is highly aggressive and is associated with an elevated mortality rate. It is challenging to distinguish VRL from uveitis because both are associated with vitreous opacity and yellow subretinal lesions^3, 4^. However, delaying diagnosis may have vision- and life-threatening consequences. The golden standard for VRL diagnosis is commonly based on clinical presentation along with histopathologic or cytologic examinations of the ocular specimens including retinal tissue and vitreous fluid (VF). The presence of malignant B lymphocytes is identified via immunohistochemistry^5, 6^. However, limited cellular yield, risk of cell lysis due to the fragile nature of lymphoma cells, and lack of experience of cytopathologists contribute to the difficulty in cytologic diagnosis^7^. Additionally, retinal tissue biopsies are invasive due to the inherent risks associated with retinal surgery^8^.

To improve the safety and accuracy of VRL diagnosis, alternative techniques based on intraocular fluid (IOF) samples are emerging. The anterior chamber paracentesis and diagnostic vitrectomy have been widely used in ophthalmic clinic for IOF sampling, which are relatively less invasive. The detection of interleukin-10 (IL-10) and IL-6 using flow cytometry as well as the detection of IGH gene rearrangements in lymphoma cells using polymerase chain reaction (PCR) and in IOF samples are supportive for VRL diagnosis. However, the sensitivity and specificity of IL-10/IL-6 ratios and gene rearrangement in different studies ranges from 60-90%, and are largely affected by sample quality, choice of primers sets, and the treatment prescribed (e.g., corticosteroids)^9-11^. Furthermore, ocular inflammatory conditions can induce oligoclonal expansion of B lymphocytes and lead to false positive diagnosis^11^. Thus, developing a detection method with high sensitivity and specificity for the diagnosis of VRL is of paramount importance.

The circulating cell-free DNA (cfDNA) represents the endogenous DNA secreted by or derived from cells and can be detected in bodily fluids or blood. The circulating tumor DNA (ctDNA) harbors the tumor-derived genetic alterations and has been recognized as a tumor-specific biomarker; it has presented encouraging results in cancer diagnosis, prognosis, as well as treatment of multiple solid cancers^12-14^. Recently, there is a growing interest in the genetic mutations that can be observed from plasma or cerebrospinal fluid (CSF) due to their potential in CNS lymphoma diagnosis and management^15, 16^. Hiemcke-Jiwa. et al. suggested that the anterior humor (AH)-based MYD88^L265P^ mutation analysis would provide an liquid biopsy tool to diagnose and monitor VRL patients^17^. However, MYD88 mutations can only be found in approximately 70% of VRL patients^18, 19^, therefore, more various disease-specific mutations should be identified for better diagnosis. Although genetic mutations have been shown to be associated with overall survival and differential response to chemotherapy in systemic DLBCL patients, tumor-related genetic mutation detection techniques based on cfDNA remain underutilized in ocular oncology. However, only few studies focused on the genetic mutation profile of VRL.

Next-generation sequencing (NGS) technology has emerged as a promising approach for cfDNA mutation profiling characterized by high-throughput, high sensitivity, and high specificity. Thus, we performed NGS by using a panel of 446 hematopoietic tumor-related genes, which was published in previous studies on lymphoma and hematologic malignancies^20-22^. The 446-gene panel was used to detect cfDNA mutations in IOF samples from a training group of diagnosed VRL and uveitis patients, and validated using IOF samples from another group. The clinical records were analyzed retrospectively, aiming to explore the clinical utility of IOF-based NGS detection for VRL diagnosis.

## Method

### Patient selection, sample collection, treatment and follow-up in the training group

Patients who were diagnosed with VRL or uveitis and had cfDNA mutation analysis performed at Zhongshan Ophthalmic Center (ZOC) of the Sun Yat-Sen University, Guangzhou, China, between April 1^st^, 2018 and March 1^st^, 2021, were selected as part of the training group. A written informed consent was collected from each patient and the study design was approved by the Ethics Committee of Zhongshan Ophthalmic Center, Sun Yat-sen University (NO.2021KYPJ190). All procedures were performed in compliance with the principles of the Helsinki Declaration.

Patients who presented with severe vitreous opacity, diffuse subretinal yellow lesions, or both were suspected of VRL by two experienced ophthalmologists (D. Liang and WR. Su) during the initial stage and got a definitive diagnosis of VRL or uveitis at a follow-up examination. To clarify the diagnosis, routine slit lamp eye examination, fundus photography, fundus fluorescein angiography (FFA), indocyanine green angiography (ICGA), optical coherence tomography (OCT), and B-ultrasound examination were performed. Laboratory investigations for autoimmune diseases or other etiology of uveitis (e.g., tests for syphilis, *Mycobacterium tuberculosis* infection, and antiviral antibodies, chest CT, urinalysis, and kidney and liver function tests) were routinely performed. Some patients underwent brain magnetic resonance imaging (MRI) or cytologic examination of CSF to exclude CNS lymphoma.

Anterior chamber paracentesis and diagnostic vitrectomy were performed to collect IOF sample from these patients as previously reported^23^. The undiluted samples were immediately transported to a cytopathologist for further analysis. The samples were added to glass slides: one slide was stained with May-Grünwald-Giemsa and the rest were used for immunostaining^24, 25^. The IOF samples were tested for the IL-10/IL-6 ratio, gene rearrangement, or cfDNA genetic mutations (either AH or VF, or both). The results of the aforementioned medical tests were reviewed for all included patients.

The therapeutic strategies were determined by the two ophthalmologists (D Liang and WR Su). Patients presenting VRL-specific clinical features that were positive for pathologic examinations or gene rearrangements, or exhibited an IL-10/IL-6 ratio >1 were treated with vitreous injection of methotrexate (MTX, 400 mg/0.1 ml). The oncologist (HQ. Huang) specialized in cerebral lymphoma was invited to perform systemic evaluations and provide the required treatment. Patients exhibiting less obvious symptoms of VRL underwent a corticosteroid treatment combined with/without immunosuppressive therapy or anti-infective treatments based on the patient’s condition. The therapeutic strategies were adjusted, and the diagnoses were given following careful evaluation of treatment effects during follow-ups.

### VRL and uveitis diagnostic criteria

Combined disease-specific ocular manifestations with the results of pathology, IL-10/IL-6 ratio, gene rearrangements and the response of patients to treatments, patients were divided into two groups: the VRL group and the uveitis group. The diagnoses were given by two independent ophthalmologists (D Liang and WR Su) independently. Any disagreements on diagnosis and treatment were resolved through discussions and, if required, referred to a third ophthalmologist (XQ Chen).

The diagnostic criteria were as follows:

VRL diagnosis was definitively established by lymphoma cells detection in pathological examinations of ocular specimens or pathologically proven systemic lymphoma, combined with disease-specific ocular manifestation and positive response to anti-tumor therapy. Patients without positive pathological evidence, presenting VRL-specific features, exhibiting a positive response to anti-tumor therapy as well as negative responses to other therapies during the follow-ups, and/or exhibiting an elevated IL-10/IL-6 ratio, and/or positive for IGH gene rearrangements, were diagnosed with VRL.

Uveitis diagnosis was definitely established following positive laboratory test results for autoimmune disease and positive responses to corticosteroids with/without immunosuppressive therapy, or positive laboratory test results for infectious uveitis and positive responses to anti-infective treatment. Furthermore, uveitis diagnosis was based on the lack of sufficient positive results from laboratory investigations including pathological examinations, IL-10/IL-6 ratio, or IGH gene rearrangements to support lymphoma diagnosis.

We classified the VRL lesions as previous reported^26^. Briefly, (1) the vitreous opacity type, exhibiting a vitreous opacity of 2+ or higher without retinal lesions; (2) the retina type, exhibiting a vitreous opacity of 1+ or less with retinal lesions only; or (3) the concomitant type, exhibiting both.

### The diagnostic value of cfDNA genetic mutations analysis for VRL

The diagnostic value including sensitivity, specificity, positive and negative predictive values, and test efficiency of genetic mutation analysis in diagnosing VRL were analyzed as previously described^27^. The detection of targeted genetic mutations was considered a positive result and recorded as ctDNA (+).

### Validation of the diagnostic value of cfDNA mutations

The VRL and uveitis validation groups were composed of patients from the Ophthalmic Unite in Beijing Chaoyang Hospital, Beijing, China. The aforementioned diagnostic criteria were used. Diagnosis was made by two experienced ophthalmologists (Yong Tao and Zhuyun Qian) independently. Any disagreements were resolved through discussions and, if required, referred to a third specialist. IOF samples from the patients from the validation set were sent for cfDNA mutation detection. All patients provided an informed consent and the institutional review board approval was obtained. The study was performed in accordance with the tenets of the Declaration of Helsinki.

### Mutation analysis by next-generation sequencing of cfDNA

#### cfDNA extraction

Samples were first transported to the central testing laboratory (Nanjing Geneseeq Technology Inc. Nanjing, China). cfDNA was extracted and tested for the 446 targeted genes. cfDNA was extracted using the QIAamp Circulating Nucleic Acid Kit (QIAGEN). Genomic DNA of the whole blood sample was prepared by DNeasy Blood & Tissue kit (QIAGEN) as control for germline mutations. DNA was quantified by dsDNA HS Assay Kit (Life Technologies, Eugene, OR, USA) according to the manufacturer’s recommendations.

#### Library Preparation

Sequencing libraries were prepared using the KAPA Hyper Prep kit (KAPA Biosystems, Cape Town, South Africa) with an optimized manufacturer’s protocol. In brief, 50ng-1μg of genomic DNA was sheared into 350 bp fragments using the Bioruptor Pico, and then underwent end-repairing, A-tailing and ligation with indexed adapters sequentially, followed by size selection using Agencourt AMPure XP beads. Finally, libraries were amplified by PCR and purified for target enrichment.

#### Hybridization Capture and Sequencing

Different libraries with unique indices were pooled together in desirable ratios for up to 2 μg of total library input. Human cot-1 DNA (Life Technologies, Carlsbad, CA, USA) and xGen Universal blocking oligos (Integrated DNA Technologies, Coralville, IA, USA) targeting 446 lyphoma-related genes were used for hybridization enrichment. The capture reaction was performed with the NimbleGen SeqCap EZ Hybridization and Wash Kit (Roche,Madison,WI,USA) and Dynabeads M-270 (Life Technologies,Vilnius,Lithuania) with optimized manufacturers’ protocols. Captured libraries were on-beads amplified with Illumina p5 and p7 primers in KAPA HiFi HotStart ReadyMix (KAPA Biosystems, Cape Town, South Africa). The post-capture amplified library was purified by Agencourt AMPure XP beads and quantified by qPCR using the KAPA Library Quantification kit (KAPA Biosystems, Cape Town, South Africa). Library fragment size was determined by the Agilent Technologies 2100 Bioanalyzer (Agilent, Santa Clara, CA, USA). Enriched libraries were sequenced on Hiseq 4000 NGS platforms (Illumina) to targeted mean coverage depths of at least 100x for whole blood control samples, and 5000x for cfDNAs.

#### Sequence Data Processing

Trimmomatic was used for FASTQ file quality control (QC). Reads from each sample were mapped to the reference sequence hg19 (Human Genome version 19) using Burrows-Wheeler Aligner (BWA-mem, v0.7.12). VarScan2 was employed for detection of somatic mutations. Annotation was performed using ANNOVAR on hg19 reference genome and 2014 versions of standard databases and functional prediction programs. Genomic fusions were identified by FACTERA with default parameters. Copy number variations (CNVs) were detected using ADTEx (http://adtex.sourceforge.net) with default parameters. An IOF or a plasma sample was defined as ctDNA-positive if any somatic mutations detected in the 446 gene panel. All mutations were confirmed by the Integrative Genomics Viewer (IGV v2.3). The CNVs of each gene in the 446 gene panel were estimated from the calculation of CNVs by Control-FREEC. The KEGG signal analysis were analyzed by KOBAS 3.0 (http://kobas.cbi.pku.edu.cn).

### Statistical analyses

Comparisons of the DNA concentration and mutations were performed using the Student ‘ s t-test for normally distributed data and Mann Whitney test for non-parametric data. Data are presented as the median and inter-quartile difference (IQR, 25% percentile, 75% percentile). P-values < 0.05 were considered statistically significant. The statistical analyses were performed using the Statistical Package for the Social Sciences (SPSS, version 19.0; IBM, Chicago, IL, USA).

## Results

### Demographic, clinical, and laboratory data of patients from the training group

A total of 23 patients had undergone genetic mutation analysis in ZOC from April 1^st^, 2018 to March 1^st^, 2021. Among the 23 patients, 17 were diagnosed with VRL while 6 were diagnosed with uveitis after clinical examination and follow-up treatment. The demographic, clinical, and laboratory data of patients from the training group are summarized in Table 1.

**Table 1.**
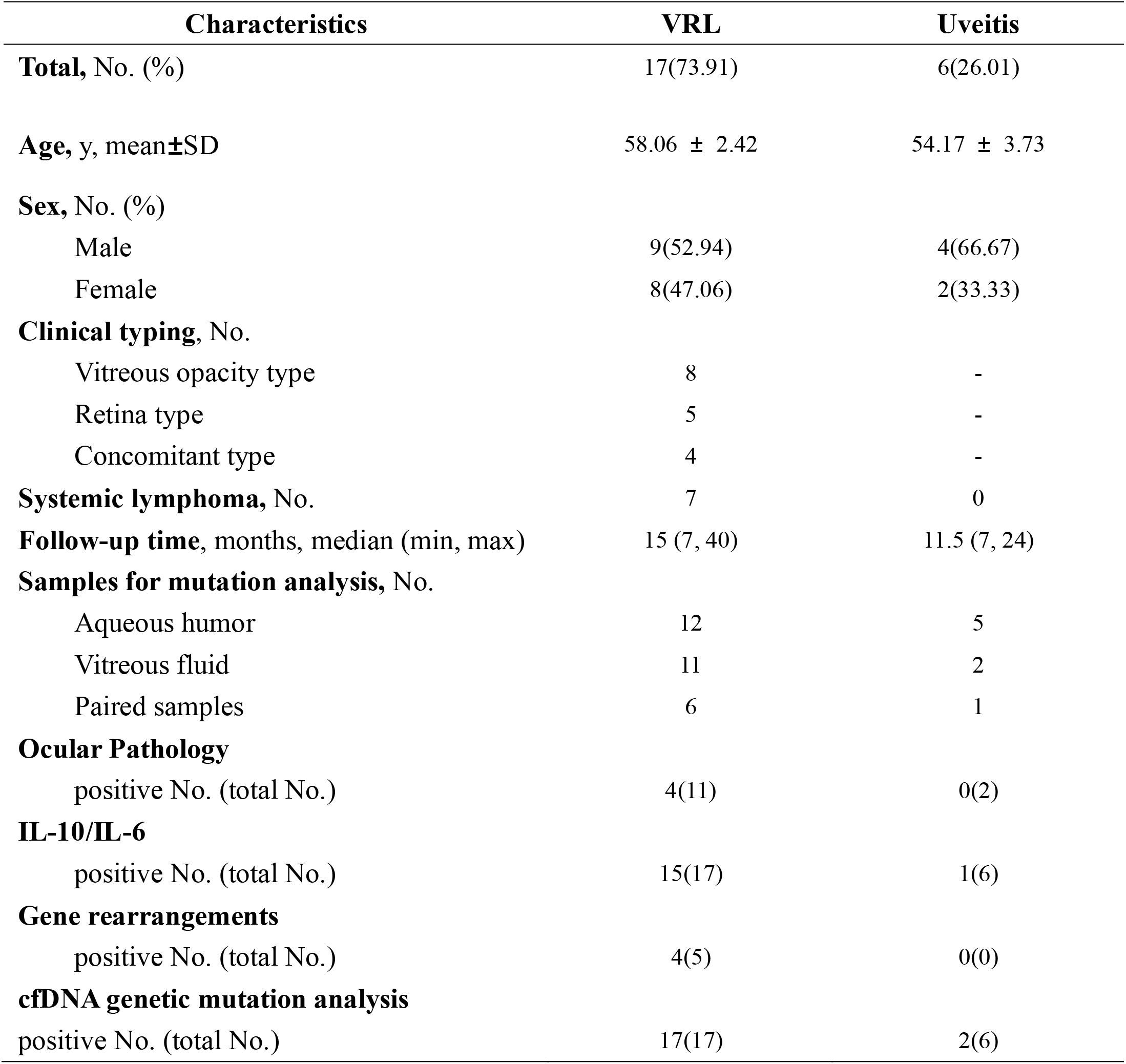
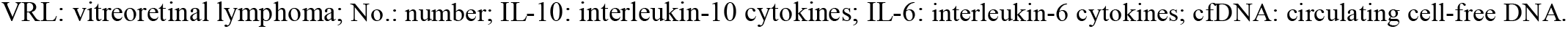
Demographic, Clinical and Laboratory Data of Patients Included in Training Group.

| Characteristics | VRL | Uveitis |
| --- | --- | --- |
| **Total, No. (%)** | 17(73.91) | 6(26.01) |
| **Age, y, mean±SD** | 58.06 ± 2.42 | 54.17 ± 3.73 |
| **Sex, No. (%)** |  |  |
| Male | 9(52.94) | 4(66.67) |
| Female | 8(47.06) | 2(33.33) |
| **Clinical typing, No.** |  |  |
| Vitreous opacity type | 8 | - |
| Retina type | 5 | - |
| Concomitant type | 4 | - |
| **Systemic lymphoma, No.** | 7 | 0 |
| **Follow-up time, months, median (min, max)** | 15 (7, 40) | 11.5 (7, 24) |
| **Samples for mutation analysis, No.** |  |  |
| Aqueous humor | 12 | 5 |
| Vitreous fluid | 11 | 2 |
| Paired samples | 6 | 1 |
| **Ocular Pathology** |  |  |
| positive No. (total No.) | 4(11) | 0(2) |
| **IL-10/IL-6** |  |  |
| positive No. (total No.) | 15(17) | 1(6) |
| **Gene rearrangements** |  |  |
| positive No. (total No.) | 4(5) | 0(0) |
| **cfDNA genetic mutation analysis** |  |  |
| positive No. (total No.) | 17(17) | 2(6) |

VRL: vitreoretinal lymphoma; No.: number; IL-10: interleukin-10 cytokines; IL-6: interleukin-6 cytokines; cfDNA: circulating cell-free DNA.

The IOF samples from the examined eye of every patient were collected before treatment and sent for diagnostic tests. Furthermore, 7 paired AH and VF samples from 7 patients were simultaneously collected by anterior chamber puncture and vitrectomy. Unpaired samples included those of which only VF or AH was available for analysis. Sixteen unpaired samples (10 AH and 6 VF) were collected from 16 patients; 22 blood samples and 9 CSF samples were also collected.

A total of 17 patients (17/23, 73.91%) were diagnosed with VRL, of which 4 (23.53%) were diagnosed through ocular pathology, and 7 (41.18%) were diagnosed by identifying the development of systemic lymphoma combined with typical ocular manifestation, and 6 (35.29%) were diagnosed by VRL-specific ocular manifestation combined with positive response to anti-tumor therapy and/or exhibiting an elevated IL-10/IL-6 ratio, and/or positive for IGH gene rearrangements. Of these VRL patients, 15 (88.24%) had an IL-10/IL-6 ratio ≥1 and responded well to anti-tumor therapy. Alternatively, 2 patients showed alleviated symptoms following MTX intravitreal injection, although the IL-10/ IL-6 ratio was <1. Five VRL patients had gene rearrangements tests, of which 4 were positive. In the uveitis group, these 6 patients (6/23, 26.09%) were proven to have different etiology, including autoimmune uveitis, virus uveitis and diffuse uveal melanocytic proliferation. One viral infuveitis patient was negative for cfDNA mutation analysis but had a very high concentration of IL-10 in IOF. The elevated IL-10 made the diagnosis confused, but the patient had symptoms alleviated after the anti-virus and glucocorticoids treatment for 6 months and got remission at present.

### Intraocular cfDNA genetic mutation analysis exhibited high sensitivity and specificity for VRL diagnosis in training group

The cfDNA of AH and VF samples from the 23 patients were performed with genetic mutation analysis by NGS and the results were analyzed. Among all the 23 enrolled patients, 19 patients got ctDNA positive, while 4 patients were ctDNA negative. All of the 17 VRL cases were ctDNA positive comparing with that 2 of 6 uveitis cases were ctDNA positive (Figure 1).

**Fig. 1.**
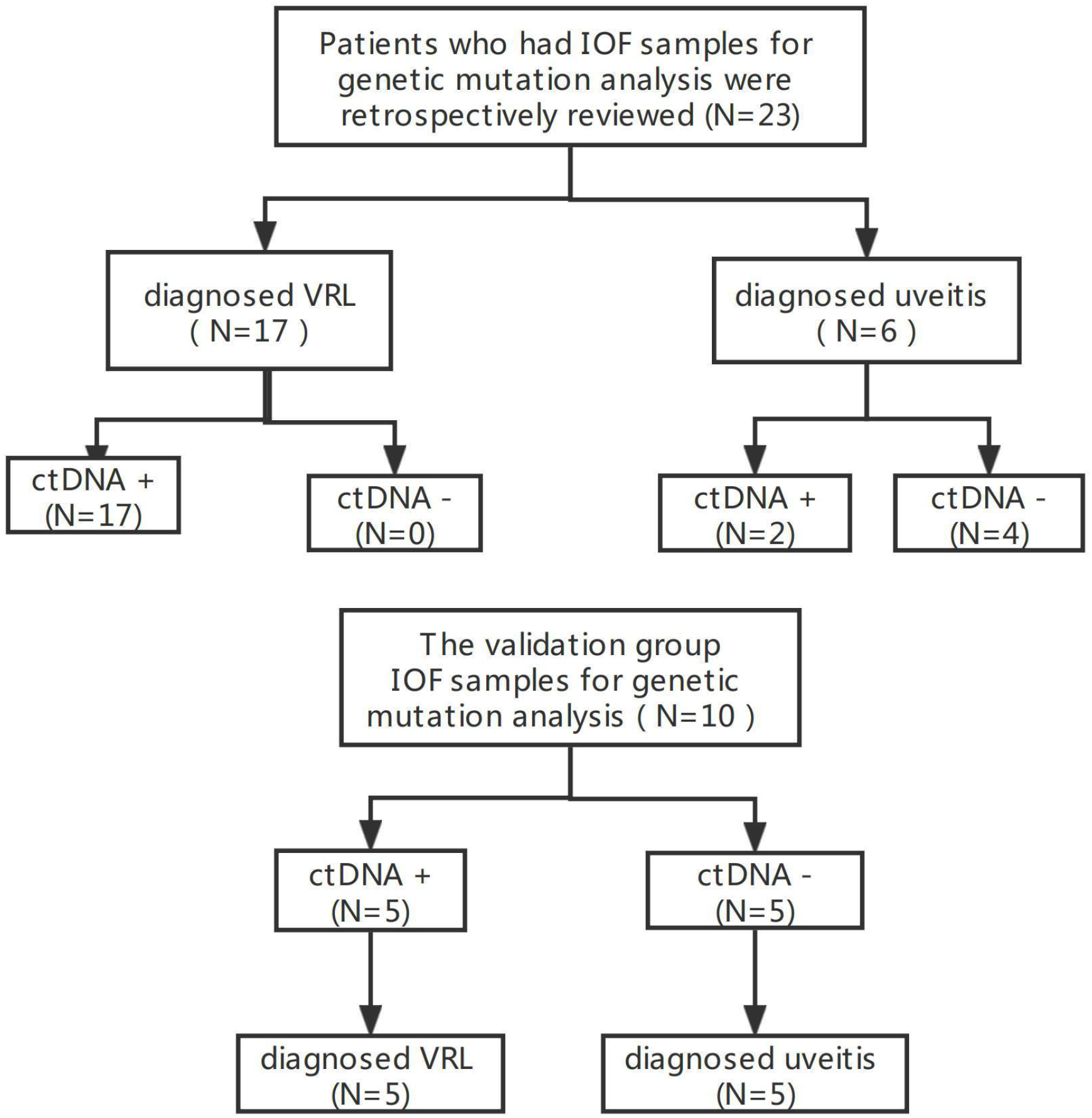
Work flow chart of patient inclusion and genetic mutation analysis.

The cfDNA concentrations observed in AH [0.48 (IQR 0.37, 0.60) ng/μl] and VH [1.5 (IQR 0.93, 4.02) ng/μl] samples from VRL patients were not significantly different from those of uveitis patients [0.38 (IQR 0.15, 0.47) in AH, 0.24 (IQR 0.20, 0.28) ng/μl in VH, respectively]. However, the ctDNA concentration, MAF mean, and number of somatic mutations observed in VRL patients were significantly higher than those observed in uveitis patients (Table 2, Figure 2).

**Table 2.** Description and Comparison of Genetic Mutation Analysis Results Between VRL and Uveitis Patients in Training Group.

|  | Anterior Humor |  |  |  | Vitreous Fluid |  |  |  |
| --- | --- | --- | --- | --- | --- | --- | --- | --- |
|  | cfDNA<br>concentration<br>(ng/μl) | ctDNA<br>concentration<br>(hGE/ml) | mean MAF | number of<br>somatic<br>mutation | cfDNA<br>concentration<br>(ng/μl) | ctDNA<br>concentration<br>(hGE/ml) | mean MAF | number of<br>somatic<br>mutation |
| **VRL** | 0.475<br>(0.3650, 0.6000) | 56040<br>(50682, 79880) | 45.5%<br>(35%, 49%) | 23<br>(13.5, 36.0) | 1.5<br>(0.9300, 4.020) | 164217<br>(108941, 336531) | 39.2%<br>(33%, 41%) | 18<br>(12.0, 24.0) |
| **Uveitis** | 0.38<br>(0.1490, 0.4740) | 0<br>(0, 32177) | 0.0%<br>(0, 8.1%) | 0<br>(0, 1) | 0.239<br>(0.1980, 0.2800) | 0<br>(0, 0) | 0<br>(0, 0) | 0<br>(0, 0) |
| **P value** | 0.1367 | 0.0092 | 0.0039 | 0.0003 | 0.1026 | 0.0256 | 0.0256 | 0.0256 |

Data are presented as the median ( IQR, 25% percentile , 75% percentile).
VRL: vitreoretinal lymphoma; cfDNA: circulating cell-free DNA; ctDNA: circulating tumor DNA; MAF: mutation allele frequency.

**Fig. 2.**
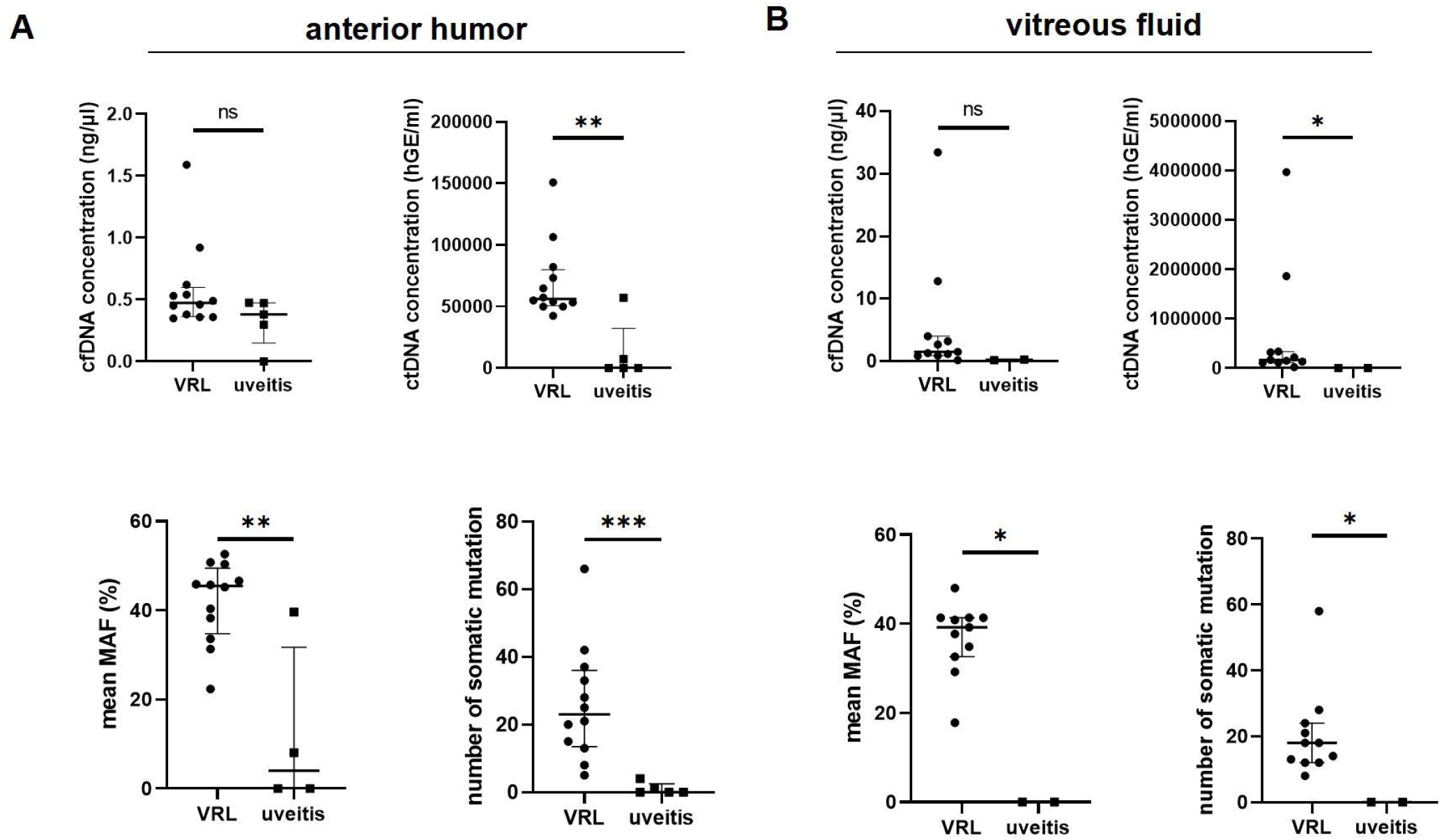
Comparison of genetic mutations observed in the anterior humor and vitreous fluid samples of VRL and uveitis patients from the training group. **(A, B)** There was no significant difference in the cfDNA concentration between VRL and uveitis patients. However, the ctDNA concentration, mean MAF and number of somatic mutations of both anterior humor and vitreous fluid samples were higher in VRL patients than in uveitis patients.

Two of the 6 uveitis patients (patient 18 and 20) tested positive for ctDNA. Patient 18 presented with severe vitreous opacity in the left eye in October 2020. He had a history of ankylosing spondylitis and uveitis for 20 years. His vision fluctuated due to irregular immune suppressive treatment. The condition improved following vitrectomy surgery and achieved remission after one year of adalimumab treatment. An *IKZF3* mutation at an MAF of 39.6% was detected in his AH sample, with a ctDNA concentration of 57,062 hGE/ml. However, the associated IL-10/IL-6 ratio was <1. Patient 20 was diagnosed with autoimmune uveitis, showed symptom alleviation after immune suppressive agent treatment, and reached a stage where the discontinuation of drugs was possible. For patient 20, 4 mutations at an MAF of 8.1% were detected within the AH sample, including *ASXL1, CHEK2, POT1*, and *FLT4* mutations. The ctDNA concentration (7,292 hGE/ml) was relatively low compared to that of the VRL patients [56,040 (IQR 50,682, 79,880) hGE/ml in AH]. Nevertheless, this patient did not exhibit elevated IL-10 level. Remarkably, the 5 mutations found in the two uveitis patients were not common in B-cell lymphomas.

The diagnostic values including sensitivity, specificity, positive and negative predictive value, and test efficiency are presented in Table 3. The genetic mutation analysis of IOF samples showed a high diagnostic potential and efficiency for VRL. Compared with the genetic mutation analysis, the sensitivity and specificity of IL-10/IL-6 ratio were 88.24% and 83.33% respectively among these 23 patients.

**Table 3.** Diagnosis Value of cfDNA Genetic Mutation Analysis for VRL Diagnosis in Training Group.

| Genetic mutations in cfDNA | VRL patients | Uveitis patients |
| --- | --- | --- |
| Positive; ctDNA(+) | 17 | 2 |
| Negative; ctDNA(-) | 0 | 4 |
| Sensitivity | 1 |  |
| Specificity | 0.67 |  |
| Positive predictive value | 0.89 |  |
| Negative predictive value | 1 |  |
| Test efficiency | 0.91 |  |

cfDNA: circulating cell-free DNA; VRL: vitreoretinal lymphoma; ctDNA: circulating tumor DNA. The detection of genetic mutations was considered as a positive result and recorded as ctDNA (+).

### Validation of cfDNA genetic mutation analysis as a diagnostic method for VRL

To consolidate our findings, 10 patients from the Beijing Chaoyang Hospital were enrolled into the validation group. A total of 10 VF samples were collected and sent for cfDNA genetic mutation analysis. Five ctDNA positive patients were VRL patients, while 5 ctDNA negative patients were uveitis patients (Figure 1). The results indicate a 100% sensitivity and specificity for VRL diagnosis.

The cfDNA concentrations, ctDNA concentration, MAF, and number of somatic mutations of all validation samples are statistically analysed and presented in Table 4 and Figure 3. Although there were no differences in the cfDNA concentration between VRL and uveitis samples (*P* = 0.310), the IOF samples from VRL patients were ctDNA positive, with a significantly higher ctDNA concentration [192,530 (IQR 110,371, 1,114,886) hGE/ml), *P* = 0.008], MAF [31.7% (IQR 27.8%,39.9%), *P* = 0.008], and number of somatic mutations [19 (IQR 7.5, 23.0), *P* = 0.008] than those observed in IOF samples from uveitis patients.

**Table 4.**
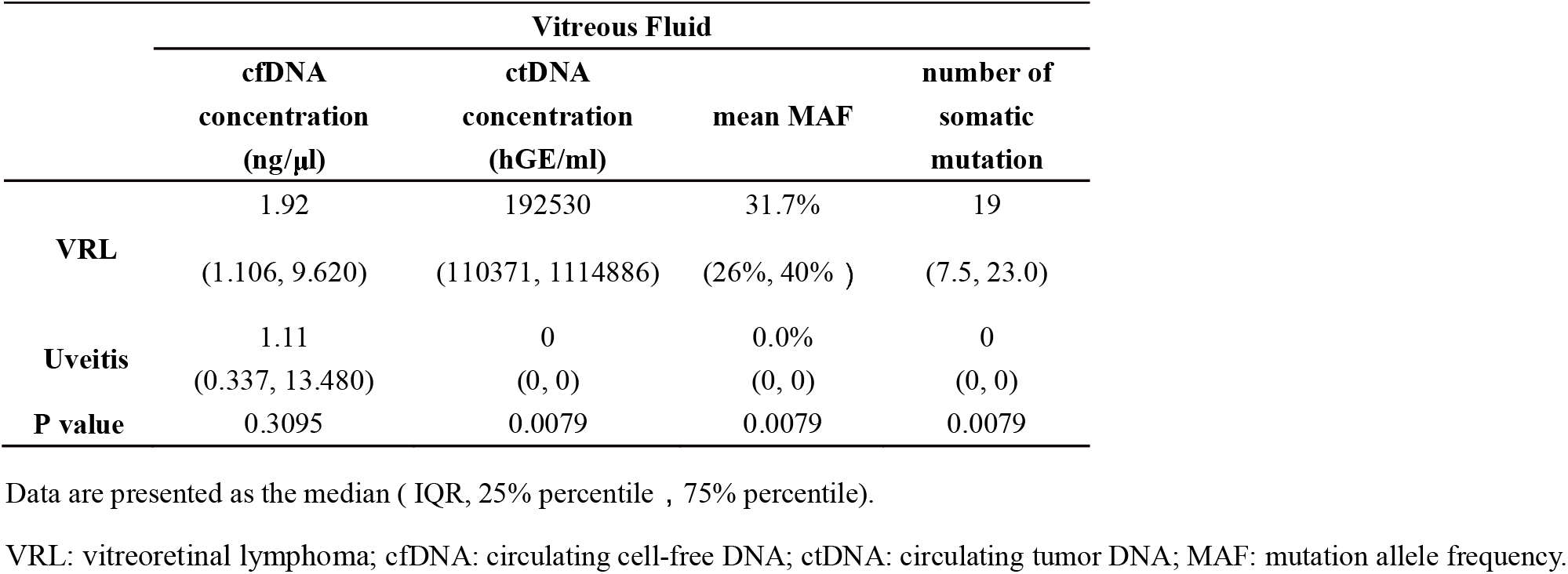
Description and Comparison of Genetic Mutation Analysis Results Between VRL and Uveitis Patients in Validation Group.

**Fig. 3.**
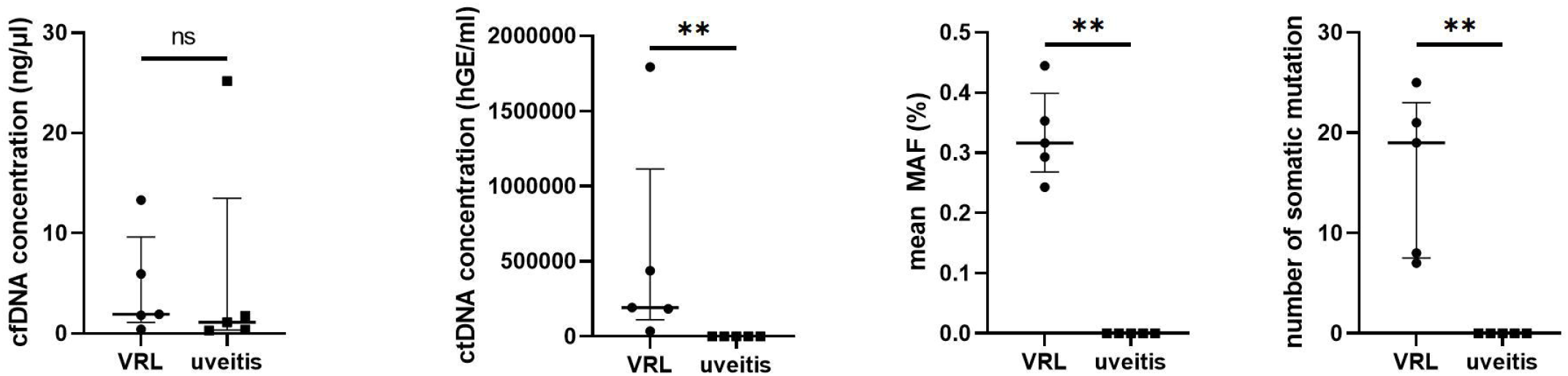
Comparison of genetic mutations observed in the vitreous fluid samples of VRL and uveitis patients from the validation group. There was no significant difference in the cfDNA concentration between VRL and uveitis patients. However, the ctDNA concentration, mean of MAF and number of somatic mutations was higher in VRL patients than in uveitis patients.

### Both AH and VF samples show similar cfDNA genetic mutations

Furthermore, we compared the genetic mutations in AH and VF samples of VRL patients to evaluate the diagnostic value of different IOF samples. cfDNA concentration (ng/μl) and ctDNA concentration (hGE/ml) were significantly higher in VF samples than in AH samples (*P* = 0.002 and 0.002). However, MAF and the number of genetic mutations were not different between AH and VF samples (Figure 4A). Furthermore, the types and frequencies of the top 12 high-frequency mutations were similar in paired samples obtained from VRL patients (Figure 4B). Among the 7 paired IOF samples, the ctDNA detection rate of AH and VF samples were the same, showing similar potential for VRL diagnosis.

**Fig. 4.**
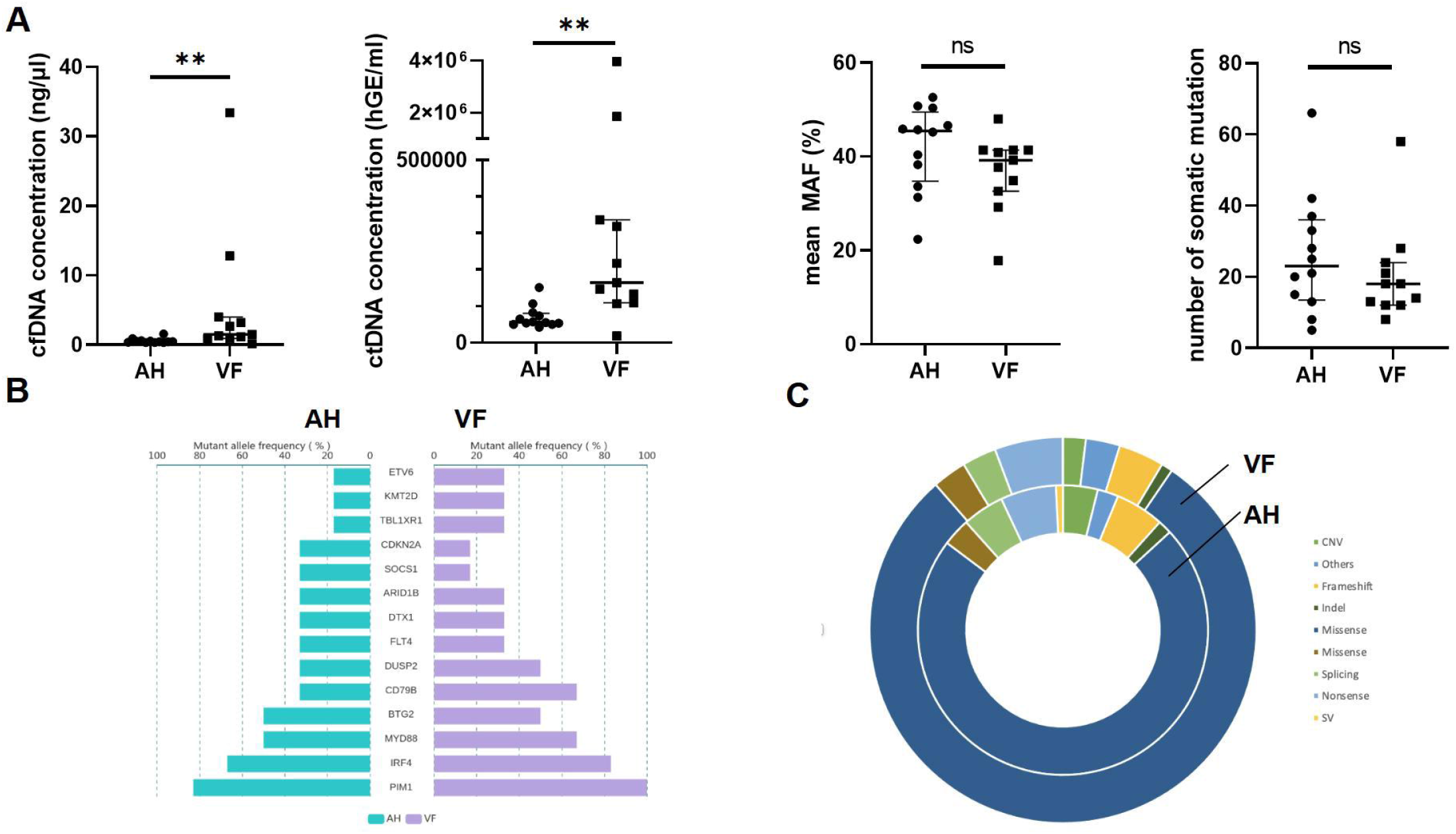
The genetic mutations observed in anterior humor and vitreous fluid samples show equal VRL diagnostic value. **(A)** The cfDNA concentration and the ctDNA concentration in VF samples were significantly higher than those in AH samples, while the mean of MAF and the number of genetic mutations showed no difference. **(B, C)** For the paired IOF samples, the frequencies of the top 12 high-frequency mutations and the types of mutations were similar in both AH and VF samples.

### Genomic mutation profiling of 22 VRL patients

All samples underwent the 446-gene panel sequencing, and the mutation profile of the 22 VRL patients (17 from training group and 5 from validation group) are illustrated using a bubble plot (Figure 5A). *PIM1* (21/22, 90.91%), *MYD88* (17/22, 77.27%), *CD79B* (11/22, 50.00%), *ETV6* (11/22, 50.00%), and *IRF4* (11/22, 50.00%) were 5 of the most frequently mutated genes found in these VRL patients.

**Fig. 5.**
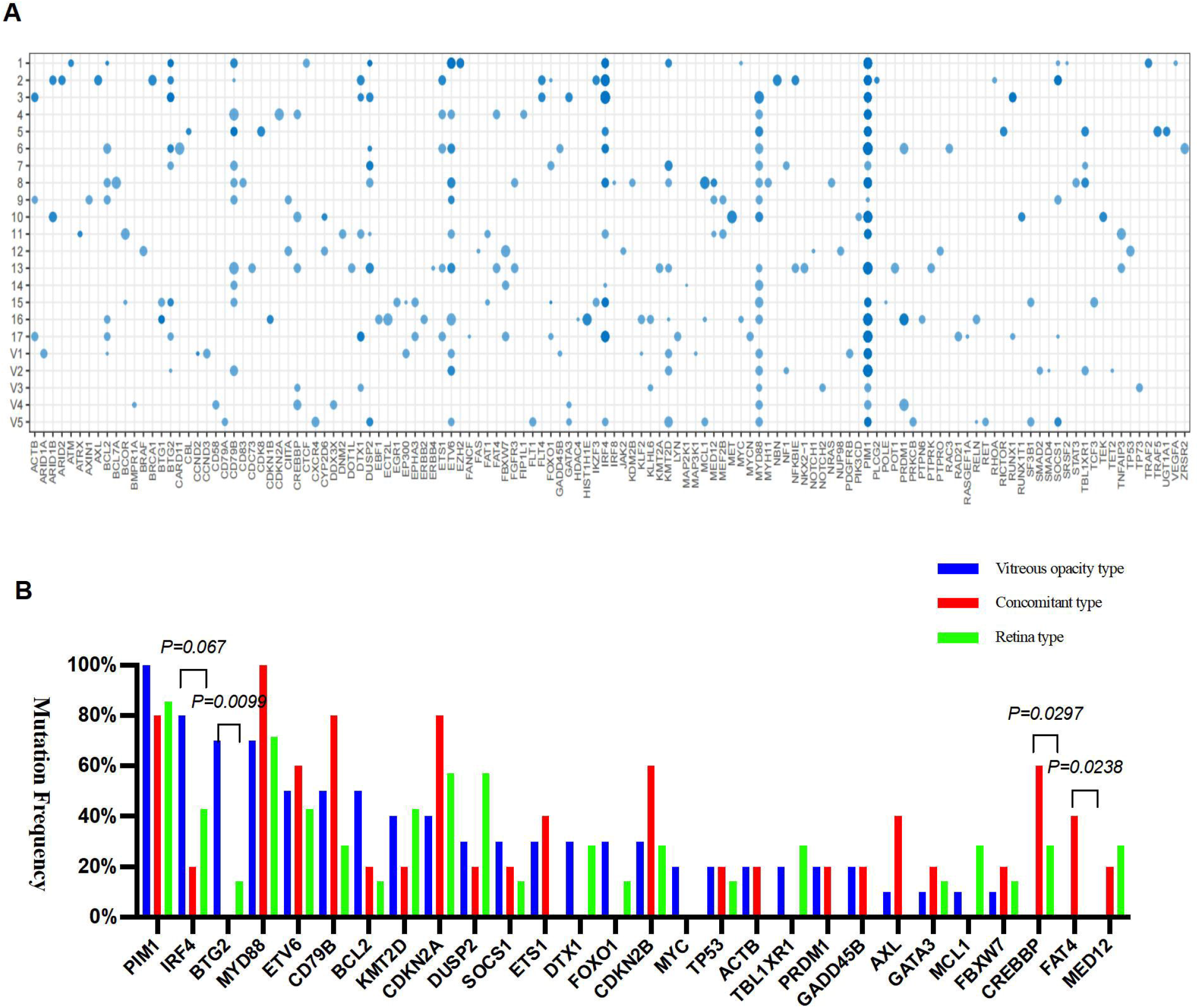
Genomic profiling of targeted mutations in 22 VRL patients. **(A)** Bubble plot representing all detectable gene mutations identified in each patient. The bubble size represents the mutation allele frequency (MAF). **(B)** The bar graph illustrates the distribution of some high-frequency mutations among the VRL patients of the vitreous opacity type, the concomitant type, and the retina type.

Among these VRL patients, 7 patients presented as retina type, 10 were vitreous opacity type and 5 were concomitant type. The mutations specific for the different subtypes were analyzed (Figure 5B). We discovered that the mutation frequency of *BTG*2 was significantly higher in the vitreous opacity type than in the retinal type (*P* = 0.0099). Furthermore, the *MYC* gene had been only detected in the vitreous opacity type, yet the *CREBBP, FAT4*, and *MED12* genes had not been detected in the vitreous opacity type. Moreover, the *ACTB, PRDM1, GADD45B*, and *ALX* genes had not been detected in the retinal type either.

### Genetic subtypes and lymphocyte differentiation pathways of genetic mutations of VRL patients

The genetic subtypes involved in different mutations of lymphocyte differentiation pathway have distinct clinical outcomes. Cell of origin (COO) classification of DLBCL has been the most commonly used subtyping method. In this study, we firstly classified the VRL patients into germinal-center B-cell-like (GCB), non-GCB, and unclassified types using the classifier described by Scherer et al^28^. In our samples, 95.5% (21/22) were non-GCB subtype, while patient 17 was unclassified (Figure 6A). Recently, George et al.^29^ established a LymphGen probabilistic algorithm to facilitate the application of DLBCL genetic subtyping, which is associated with the response to chemotherapy. By using this algorithm, we determined the genetic subtype of each VRL patient. The nosology covered 86.36% (19/22) of VRL patients, of which 17 (77.27%) were classified as the MCD subtype and 2 (9.09%) were classified as the BN2 subtype (Figure 6B). Patient 17, who was unclassified based on the COO classification, was classified as BN2 subtype by the LymphGen algorithm with a 0.76 confidence.

**Fig. 6.**
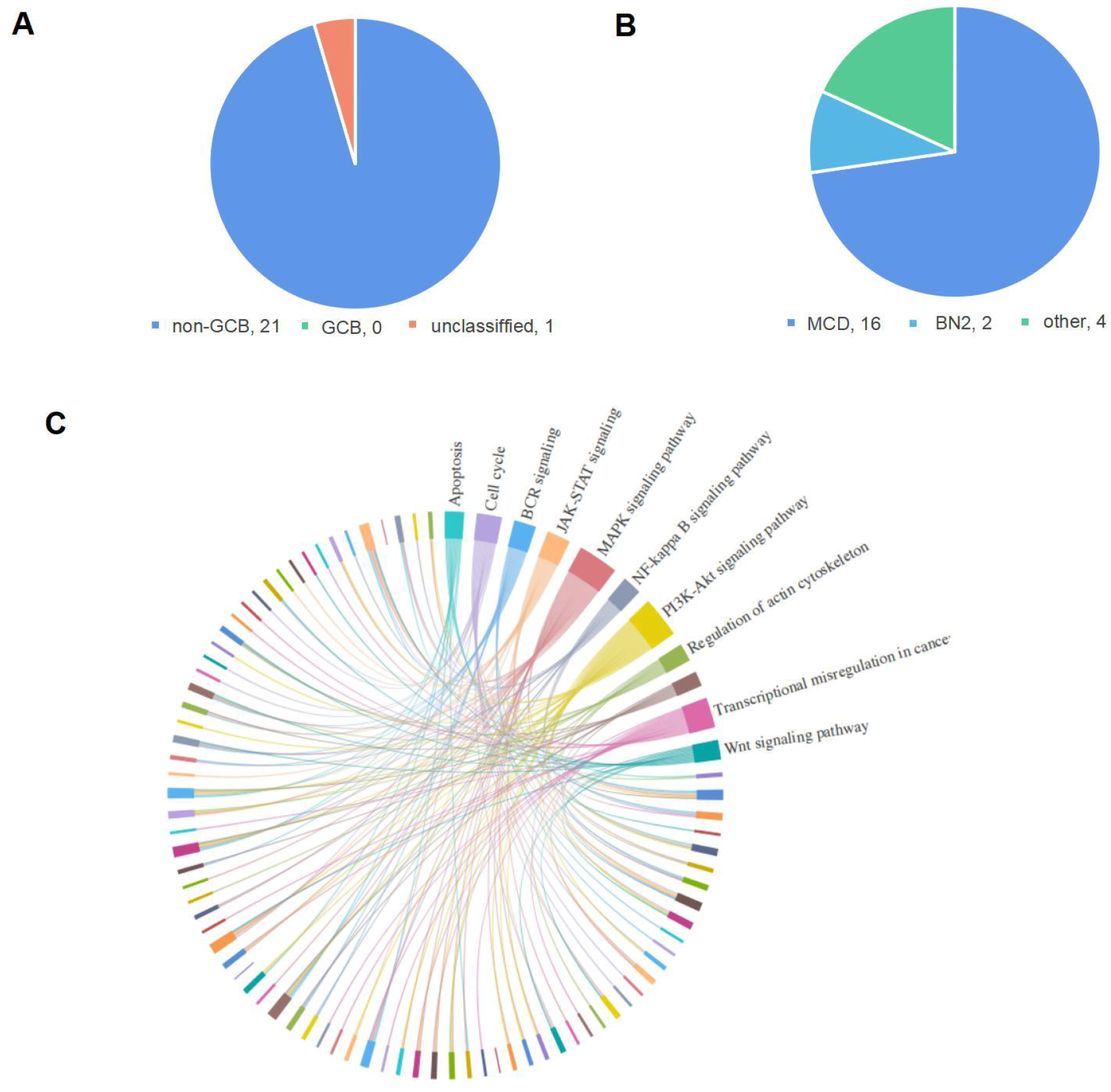
Subtype prediction and lymphocyte differentiation pathways of genetic mutations among VRL patients. **(A, B)** The subtypes of VRL patients based on CCO (cell of origin) classification. The majority of VRL patients (95.45%) in our study were non-GCB type. Further classification using the LymphGen probabilistic algorithm: 17 VRL patients were classified as the MCD subtype, and 2 patients were classified as the BN2 subtype. **(C)** The genetic mutations of VRL patients were divided into specific pathways that were associated with B-cell differentiation. The signaling pathways that were involved and prevalent in VRL patients are illustrated.

At the same time, we divided the genetic mutations of VRL into specific pathways that were associated with B-cell differentiation. It was demonstrated that the VRL patients in our study were mostly characterized by mutations in the PI3K-Akt signaling pathway, followed by MAPK, transcriptional mis-regulation in cancer, Ras, JAK-STAT, and NF-kB signaling pathways (Figure 6C).

## Discussion

VRL, a DLBCL non-Hodgkin’s lymphoma, is an aggressive type of cancer with a high mortality rate^1^. The pathogenesis of VRL is largely unknown and its diagnosis remains a challenge for clinicians. Liquid biopsy, used for the detection of ctDNA has been shown to represent a valuable diagnostic method^14, 15^. However, it is underutilized in VRL diagnosis. In our study, we explored the diagnostic value of genetic mutation analysis of IOF samples using NGS and a panel of 446 tumor-related genes for VRL diagnosis. In the training group, all VRL patients tested positive for mutations (sensitivity 100%), while only 2 out of the 6 uveitis patients were positive (specificity 67%). However, the mutations detected in uveitis patients were identified as clonal hematopoiesis mutations. In the validation group, both the sensitivity and specificity of cfDNA genetic mutation analysis for VRL diagnosis were 100%. Furthermore, we showed that the AH and VF samples had similar VRL diagnostic values when used for disease-specific mutation detection. Additionally, we discovered that the top five mutated genes in VRL patients were *PIM1* (21/22, 90.91%), *MYD88* (77.27%), *CD79B* (50.00%), *ETV6* (50.00%), and *IRF4* (50.00%). The altered genes are primarily involved in the PI3K-Akt signaling pathway. Among the 22 VRL patients, 21 (95.45%) were classified as non-GCB while 1 was an unclassified subtype. Using the LymphGen probabilistic algorithm, 77.27% of VRL patients were classified as MCD subtype, and 2 patients (9.1%) were classified as BN2 subtype.

Numerous cfDNA mutation were detected in IOF samples from all VRL patients, which ultimately indicated an increased possibility of the presence of tumor cells (100% sensitivity). Previous studies introduced additional liquid biopsy tools to detect mutations and support VRL diagnosis, such as MYD88^L265P[7]^. Unfortunately, the sensitivity of these tools was not satisfactory because the types of mutations they tested were limited. The panel of 446 tumor-related genes used here has an advantage in tracking novel or rare mutations in VRL, thus reaching a 100% sensitivity for VRL diagnosis. In addition, the diagnostic value of genetic mutations of AH and VF samples were compared. We showed that although the concentration of cfDNA and ctDNA were significantly higher (*P* = 0.002 and 0.002, respectively) in the VF samples than in the AH samples, the MAF, number of genetic mutations, and types as well as frequencies of the top 12 high-frequency mutations were similar. Considering the safety and simplicity of anterior chamber paracentesis, we suggest AH sampling as the feasible method to collect biopsy samples meant to undergo genetic mutation analysis and aid VRL diagnosis clinically.

We observed that 2 AH samples from 6 uveitis patients of the training group were positive for the genetic mutation analysis. However, the detected mutant genes (*IKZF3, ASXL1, CHEK2, POT1*, and *FLT4*) were not common with those observed in B-cell lymphoma and were considered to be clonal hematopoietic mutations^30-32^. Clonal hematopoiesis is a common aging-associated process characterized by the accumulation of leukemia-associated mutations in the hematopoietic stem cells^33^. Thus, performing a comprehensive analysis of the clinical characteristics, concentrations and numbers of mutations, as well as specificity of the detected mutated genes when getting a positive result is essential. The detection of numerous high-frequency mutations, especially the reported genes derived from VRL patients, represents a great advantage for VRL diagnosis. Nevertheless, we need a higher sample size to optimize and establish a more specific panel of VRL-related genes for VRL diagnosis. Another important issue, because this was the first diagnostic accuracy study using a genetic mutation analysis and a tumor-related gene panel to diagnose VRL, we set the cut-off of mutations number to 0, which contributed to the false positive rate in the training group. Thus, a perspective study exploring the optimal cut-off mutation number for VRL diagnosis would complement our work.

In the training group, the IL-10/IL-6 ratio observed in IOF samples was examined in all patients, while the IGH gene rearrangement was tested only in 5 VRL patients. The results showed that the sensitivity and specificity of the IL-10/IL-6 ratio were 88.24% (15/17) and 83.33% (1/6), respectively, while the sensitivity of the IGH gene rearrangement was 80% (4/5). These results are consistent with those of previous reports^9-11^. Nevertheless, the IL-10/IL-6 ratio and IGH gene rearrangement sensitivities were lower than that of the genetic mutation analysis in our study.

The pathological features and molecular classification of lymphoma have potential clinical utility on lymphoma treatment and prognosis. In our study, the VRL patients were characterized by high mutation frequencies in *PIM1, MYD88, CD79B, ETV6*, and *IRF4*. Based on their genomic abnormalities, 21 patients (95.45%) were classified as non-GCB subtype, which is consistent with previous findings indicating that the non-GCB subtype accounts for most CNS lymphoma cases^34, 35^. In 2018, Schmitz et al. extended the systemic DLBCL into MCD, BN2, N1 and EZB subtypes, which had been proved to be associated with the response to chemotherapy^36, 37^. For example, it was reported that the viability of MCD cells was likely sustained by BCL2; thus, treatment targeting BCL2 may be effective against the MCD subtype. BN2 showed favorable survival compared to that of MCD in DLBCL^29^. However, the genetic molecular classification and targeted therapy of VRL is understudied. Here, 77.27% of VRL patients were classified as MCD subtype, and 2 patients (9.1%) were classified as BN2 subtype. Furthermore, we presented the genetic mutation profile and revealed the molecular characteristics of VRL, while the responses to chemotherapy and prognosis of these VRL patients is under further observation. Ultimately, our findings might provide great value for determining the diagnosis and treatment strategy for VRL patients.

In conclusion, we showed that the cfDNA genetic mutation analysis of IOF samples using a 446-gene panel represents a feasible method for VRL diagnosis. The 446 genes in our panel were comprehensive enough to provide 100% sensitivity for VRL diagnosis. However, the large number of genes contained some unspecific genes, contributing to the 67% specificity, which calls for further optimizations and a more specific panel of VRL-specific genes. Furthermore, our analysis could track the genetic profile of the disease, which revealed the genetic heterogeneity and molecular characteristics of VRL. This may contribute to the development of targeted treatments in future.

## Data Availability

All data produced in the present study are available upon reasonable request to the authors

## Abbreviations used in this article

VRL: Vitreoretinal lymphoma
IOF: intraocular fluid
cfDNA: circulating cell-free DNA
CNS: central nervous system
DLBCL: diffuse large B-cell lymphoma
VF: vitreous fluid
IL-10: interleukin-10
IL-6: interleukin-6
PCR: polymerase chain reaction
ctDNA: circulating tumor DNA
CSF: cerebrospinal fluid
AH: anterior humor
NGS: Next-generation sequencing
FFA: fundus fluorescein angiography
ICGA: indocyanine green angiography
OCT: optical coherence tomography
MRI: magnetic resonance imaging
MTX: Methotrexate
MAF: mutation allele frequency
GCB: germinal-center B-cell-like

## Acknowledgments

YH, XC, and DL were responsible for the study design, data collection, data analysis, and manuscript writing. XC, YH, WS, SY, XW, PZ, ZQ, HH, YT, and DL contributed to the patients’ follow-up and clinical data collection. WS, YT, HH, and DL provided guidance throughout the study. XW, PZ, XH, CL, HH, and YT helped conducting the experiment. XH, CL, ZQ, and ZL helped during data analysis and manuscript writing. HH, YT, and DL were responsible for the study design, revision of the manuscript, and final manuscript approval. All authors contributed to the article and approved the submitted version.

